# The public health impact of poor sleep on severe COVID-19, influenza and upper respiratory infections

**DOI:** 10.1101/2022.02.16.22271055

**Authors:** Samuel E. Jones, Fahrisa I. Maisha, Satu J. Strausz, Brian E. Cade, Anniina M. Tervi, Viola Helaakoski, Martin E. Broberg, Vilma Lammi, FinnGen, Jacqueline M. Lane, Susan Redline, Richa Saxena, Hanna M. Ollila

**Affiliations:** Institute for Molecular Medicine, HiLIFE, University of Helsinki, Finland; Department of Immunology and Dermatology, Yale University, School of Medicine, New Haven, Connecticut, USA; Division of Sleep and Circadian Disorders, Brigham and Women’s Hospital and Harvard Medical School, Boston, MA, USA; Broad Institute of MIT and Harvard, Cambridge, MA, USA; Center for Genomic Medicine, Massachusetts General Hospital, Boston, MA, USA; Harvard T.H. Chan School of Public Health, Harvard University, Boston, MA, USA; Anesthesia, Critical Care, and Pain Medicine, Massachusetts General Hospital and Harvard Medical School, Boston, MA, USA

## Abstract

**Background:** Poor sleep is associated with an increased risk of infections and all-cause mortality, and acute sleep loss and disruption have been linked with inflammation and poorer immune control. Previous studies, however, have been unable to evidence causality between the chronic effects of poor sleep and respiratory infection risk. In light of the ongoing COVID-19 pandemic and potential future disease outbreaks, understanding the risk factors for these infections is of great importance.

**Aim:** Our goal was to understand if chronic poor sleep could be identified as a causal risk factor for respiratory infections including influenza, upper respiratory infections and COVID-19.

**Methods:** We used population cohorts from the UK Biobank (N ≈ 231,000) and FinnGen (N ≈ 327,000) with ICD-10 based electronic health records and obtained diagnoses of insomnia, influenza and upper respiratory infections (URIs) from primary care and hospital settings. We computed logistic regression to assess association between poor sleep and infections, disease free survival hazard ratios, and used summary statistics from genome-wide association studies of insomnia, influenza, URI and COVID-19 to perform Mendelian randomization analyses and assess causality.

**Findings:** Utilizing 23 years of registry data and follow-up, we saw that insomnia diagnosis associated with increased risk for infections in FinnGen and in UK Biobank (FinnGen influenza HR = 5.32 [4.09, 6.92], P = 1.02×10^−35^, UK Biobank influenza HR = 1.54 [1.37, 1.73], P = 2.49×10^−13^). Mendelian randomization indicated that insomnia causally predisposed to influenza (OR = 1.59, P = 6.23×10^−4^), upper respiratory infections (OR = 1.71, P = 7.60×10^−13^), COVID-19 infection (OR = 1.08, P = 0.037) and risk of hospitalization from COVID-19 (OR = 1.47, P = 4.96×10^−5^).

**Conclusions:** Our findings indicate that chronic poor sleep is a causal risk factor for contracting respiratory infections, and in addition contributes to the severity of respiratory infections. These findings highlight the role of sleep in maintaining sufficient immune response against pathogens as suggested by earlier work. As the current COVID-19 pandemic has increased the number of people suffering from poor sleep, safe interventions such as sleep management and treating individuals with insomnia could be promoted to reduce infections and save lives.

## Introduction

Insomnia is a condition characterized as the “persistent difficulty with sleep initiation, duration, consolidation or quality.”^1^ Insomnia is a common condition worldwide, with between 9 and 25% of the population suffering from this condition at any one time^2–4^, depending on the definition used and population studied. In the USA, this has led to the recognition of insomnia as a critical public health concern (https://www.cdc.gov/sleep/about_us.html) and as an important intervention target in future clinical studies and public health policy^5^.

Many public health studies on the negative impacts of poor sleep have been small-scale epidemiological and intervention studies that have focused on the acute, or short-term, effects of sleep disruption. These studies have indicated that acute sleep loss and sleep disruption are associated with inflammation^6^ and a greater risk of viral infection^7,8^. Additionally, a systematic review and meta-analysis of 72 studies demonstrated that acute sleep disruption was associated with increase in circulating proinflammatory cytokine IL-6 and C-reactive protein (CRP)^9^, both of which are common indicators for inflammation. In addition, it has been suggested that an acute lack of sleep would dampen or delay the development of vaccination response^10–12^, an indication that, at least in the short-term, lack of sleep may have concrete effects on the immune system and consequently on our ability to fight off infections. However, the acute effects may be transient, especially if environmentally driven, and may not typify the kind of mechanisms driven by long-term sleep disruption.

In contrast, large cohort studies of poor sleep have shown that chronic sleep loss and insomnia are associated with increased rates of both all-cause mortality^13,14^ and viral infection^15^. More recently, a review of nine small-scale studies that assessed the effect of chronic short and long sleep on risk of developing respiratory infections^16^, found that short sleep was associated with an increased overall risk of respiratory infections (HR = 1.30 [1.19, 1.42], P<1×10^−5^). Evidence collected from a number of cross-sectional studies also points to insomnia being associated with an increased prevalence of respiratory infections^17^, though cause and effect is unclear. It has been proposed that long-term insomnia leads to chronic inflammation, resulting in poorer immune response^18^ and thus an increased risk and severity of infection. However, insomnia has not yet been ascribed a causal role in respiratory infection risk due, in part, to the complex bidirectional relationship between sleep and immune function.

The effect of the ongoing COVID-19 pandemic on sleep has been well documented. A subset of individuals suffer from worse sleep, nightmares and changes in circadian rhythms^19–21^, due to the effect of quarantines, changing work and life patterns and increased stress levels, amongst other things. Sleep disruption is also a common sequela of SARS-CoV-2 infection, with a recent meta-analysis of 66 studies reporting that sleep disturbances, including post-viral insomnia, were a common feature reported by those individual studies that had collected sleep data, with some studies reporting that COVID-19 severity was a predictor for sleep disruption^22^. What is less clear, however, is the effect of pre-infection sleep disruption on the risk of developing COVID-19 and subsequent severity of the infection. One study proposed that an observed increase in the rate at which shift workers are diagnosed with COVID-19^23^ is due to greater levels of sleep and circadian disruption in this subset of the population^24^, though it isn’t possible to separate the effect of disrupted sleep rhythms from other confounders in such studies. It is therefore critically important to assess the causal effect of poor sleep on infections and especially on respiratory infections like influenza or COVID-19, as poor sleep may accentuate the effect of pandemics on public health and so could offer an intervention to reduce infection risk.

Motivated, in part, by the ongoing SARS-CoV-2 pandemic, our aim was to assess if chronic insomnia increases the risk for respiratory infections including upper respiratory tract infections and the known severe pathogens influenza and SARS-CoV-2. We first tested the hypothesis that insomnia is a risk factor for influenza, upper respiratory infections and COVID-19 using longitudinal data from over 558,000 study participants across two biobanks. We employed methods from genetic epidemiology to infer the one-directional causal associations of sleep disruption on influenza, upper respiratory tract infections and COVID-19 susceptibility, severity and hospitalization, using data from recent large-scale genetic studies of these conditions. We show that insomnia increases the risk for COVID-19, influenza, and upper respiratory tract infections.

## Results

### Survival analysis in population cohorts

To understand whether there is a discernible impact of poor sleep on subsequent risk of respiratory infection, we first performed survival analysis by testing the associations between insomnia and respiratory infections and computing multivariable adjusted hazard ratios in over 327,000 individuals from FinnGen free of relevant respiratory infectious diseases (Table 1a and Supp Table 1). FinnGen had hospital record (inpatient and outpatient) information on diagnosis of insomnia, upper respiratory infections (URI), and influenza, with up to 23 years follow-up from 1998 to 2020. We found that a prior hospital diagnosis of insomnia almost tripled the risk of a later URI diagnosis (HR = 2.83 [2.40, 3.34], P = 5.66×10^−35^) and quintupled the risk of a subsequent influenza diagnosis (HR = 5.32 [4.09, 6.92], P = 1.02×10^−35^). COVID-19 data was examined from February 2020 to May 2021, but as the pandemic occurred after the end of available records for other diagnoses we instead performed a logistic regression to test whether those with prior diagnoses of insomnia were over-represented in COVID-19 patients (Table 1a and Supp Table 2). Our analyses indicated no significant change in risk of COVID-19 infection for those previously diagnosed with insomnia (P = 0.973).

**Table 1a.** FinnGen endpoint-to-endpoint survival and logistic regression analyses results for insomnia exposure.

|  | Disease free survival analysis<br>(up to 23 years of follow-up) |  |  | Logistic Regression |  |  |
| --- | --- | --- | --- | --- | --- | --- |
| Outcome | Hazard Ratio (HR) | HR 95% CI | P | OR | OR 95% CI | P |
| Influenza | 5.32 | [4.09,6.92] | $1.02 \times 10^{-35}$ | 2.48 | [2.02, 3.02] | $1.10 \times 10^{-18}$ |
| Upper Respiratory Infection (URI) | 2.83 | [2.40, 3.34] | $5.66 \times 10^{-35}$ | 1.81 | [1.60, 2.04] | $1.39 \times 10^{-21}$ |
| COVID-19 |  |  |  | 1.24 | [0.64, 1.46] | 0.973 |

To replicate these results, we assessed the same endpoints in approximately 231,000 UK Biobank participants using hospital and primary care records collected between March 2002 and August 2019 (Table 1b and Supp Table 2). We observed modest infection risk, with survival analyses suggesting that a prior diagnosis of insomnia increased the risk of URI by 52% (HR = 1.52 [1.43, 1.61], P = 1.72×10^−45^) and increased the risk of influenza by 54% (HR = 1.54 [1.37, 1.73], P = 2.49×10^−13^) (Table 1b). As with FinnGen, the period in which both primary care and hospital records were available, in the UK Biobank, did not overlap with the COVID-19 diagnosis interval, so we were unable to assess COVID-19 infection risk using the survival analysis framework. Logistic regression, however, did not identify significant enrichment of diagnosed insomnia sufferers within COVID-positive patients (OR=1.21 [0.82, 1.70], P = 0.311) (Table 1b and Supp Table 2).

**Table 1b.** UK Biobank endpoint-to-endpoint survival and logistic regression analyses results for Insomnia exposure.

|  | Disease free survival analysis<br>(up to 17 years of follow-up) |  |  | Logistic Regression |  |  |
| --- | --- | --- | --- | --- | --- | --- |
| Outcome | Hazard Ratio (HR) | HR 95% CI | P | OR | OR 95% CI | P |
| Influenza | 1.54 | [1.37, 1.73] | $2.49 \times 10^{-13}$ | 2.12 | [1.98, 2.27] | $3.84 \times 10^{-98}$ |
| Upper Respiratory Infection (URI) | 1.52 | [1.43, 1.61] | $1.72 \times 10^{-45}$ | 2.13 | [2.04, 2.23] | $9.13 \times 10^{-247}$ |
| COVID-19 |  |  |  | 1.21 | [0.82, 1.70] | 0.311 |

### Mendelian randomization analysis

To supplement the findings of the longitudinal association analyses, we used information of the genetic associations of insomnia and respiratory infection outcomes to perform two-sample Mendelian randomization analyses, using the TwoSampleMR R package^25,26^. Combining data from a recent large-scale insomnia genome-wide association study (GWAS) meta-analysis of 2.3 million individuals^27^, a new global meta-analysis into genetic factors predisposing to COVID-19 by the COVID-19 Human Genetics Initiative^28^ and data from GWAS of URI and influenza in release 7 of FinnGen, we estimated the causal impact of insomnia on COVID-19, URI and influenza (Table 2). We identified that insomnia was causally associated with an increased risk of severe COVID-19 symptoms (inverse-variance weighted (IVW) OR = 1.64 [1.22-2.21], P = 1.02×10^−3^) and greater risk of hospitalization from COVID-19 (IVW OR = 1.47 [1.22-1.77], P = 4.96×10^−5^), a proxy for severity of infection. There was also suggestive evidence that insomnia increased the risk of initial COVID-19 infection (IVW OR = 1.08 [1.004-1.157], P = 0.037), but this did not pass the multiple testing significance threshold and the additional MR tests were non-significant, despite broadly similar causal effect estimates. We also found insomnia to be causally associated with increased risk of influenza infection (IVW OR = 1.59 [1.22, 2.07], P = 6.24×10^−4^) and URI (IVW OR = 1.71 [1.48, 1.98], P = 7.60×10^−13^). In the MR sensitivity analyses (weighted median and MR Egger analyses), in which we apply methods that are robust to pleiotropy but statistically less powerful, there was potential evidence of directional pleiotropy in insomnia’s effect on both COVID-19 severity and hospitalization risk, with the MR Egger intercept (an estimate of the total pleiotropic effect) being non-zero (Egger intercept P = 0.013 and 0.019 for severe symptoms and hospitalization risk, respectively). This suggests that some of the insomnia instruments may not be affecting COVID-19 severity and hospitalization risk directly through insomnia but via other, as yet, unknown pathways. We did not, however, see strong evidence of pleiotropy for COVID susceptibility, URI or influenza (Supp Table 3).

**Table 2.** Causal analysis results of insomnia, short sleep and a measure of sleep fragmentation of COVID severity, susceptibility and hospitalization risk, upper respiratory infection and influenza.

|  |  |  | IVW |  |  |  |
| --- | --- | --- | --- | --- | --- | --- |
| Exposure | Outcome | NVar | logOR | SE | P | Power |
| **Insomnia** | **Severe COVID** | **464** | **0.496** | **0.151** | **0.001** | **0.996** |
|  | **Hospitalized COVID** | **489** | **0.387** | **0.095** | **4.96x10<sup>-5</sup>** | **1** |
|  | *COVID Infection* | *452* | *0.075* | *0.036* | *0.037* | *0.612* |
|  | **URI** | **466** | **0.538** | **0.075** | **7.60x10<sup>-13</sup>** | **1** |
|  | **Influenza** | **466** | **0.463** | **0.135** | **0.001** | **0.959** |
| **Short Sleep** | Severe COVID | 24 | 0.208 | 0.131 | 0.113 | 0.121 |
|  | Hospitalized COVID | 24 | 0.154 | 0.091 | 0.09 | 0.145 |
|  | *COVID Infection* | *25* | *0.078* | *0.036* | *0.032* | *0.162* |
|  | URI | 24 | 0.091 | 0.056 | 0.104 | 0.081 |
|  | Influenza | 24 | 0.18 | 0.143 | 0.208 | 0.085 |
| **No. of sleep episodes** | Severe COVID | 21 | 0.065 | 0.174 | 0.708 | 0.456 |
|  | Hospitalized COVID | 21 | 0.045 | 0.111 | 0.688 | 0.086 |
|  | COVID Infection | 21 | -0.022 | 0.034 | 0.525 | 0.092 |
|  | URI | 19 | 0.037 | 0.114 | 0.746 | 0.072 |
|  | Influenza | 19 | -0.049 | 0.154 | 0.752 | 0.053 |

Rows with results in bold font are statistically significant after Bonferroni correction and those in italics are significant (IVW P<0.05) at the level of a single test but not after Bonferroni correction. NVar = number of exposure genetic instruments used.

MR also allows us to explore the causal effect of non-disease phenotypes that are not routinely captured in longitudinal health records. Habitual short sleep (typically defined as sleeping < 6 hours per night) is a common consequence, and thus a frequently observed comorbidity, of insomnia. To understand whether loss of sleep is an important factor in insomnia’s causal associations, we tested the effect of genetically instrumented short sleep, using 27 variants associated with short sleep^29^. We found suggestive evidence that habitually short sleeping increased the risk of COVID-19 infection (IVW P = 0.03), but no strong evidence that risk of hospitalization with COVID-19 was affected (IVW P = 0.08). Similarly, there was no strong causal evidence that habitual short sleep leads to an elevated risk of infection for influenza and URI (P = 0.153, and P = 0.158, respectively). As with all statistical tests, a negative finding in MR analyses could be indicative of either no true association or lack of statistical power to detect causal effects. We therefore calculated the available power to detect the causal effects we identified (Table 2) and found that habitual short sleep was generally underpowered to detect effects of those magnitudes, meaning that we can’t definitively rule out a true association. At 80% power (a well-accepted power threshold), however, we would only be able to reliably detect causal OR of >2.1 and >1.6 for COVID-19 severity and hospitalization and >1.25 for COVID-19 susceptibility, and ORs of >2.35 and >1.57 for influenza and URI respectively (Supp Table 4). If the true causal effect of short sleeping on the outcomes were larger than these thresholds, we would likely have power to detect them.

In addition to loss of sleep, increasingly fragmented sleep is a common result of insomnia. We therefore also assessed the causal impact of sleep fragmentation on the same respiratory infection outcomes using 21 genetic variants robustly associated with the number of distinct sleep “episodes” within the primary sleep period^30^, a proxy for sleep fragmentation. We did not identify a causal association of fragmented sleep on COVID-19 susceptibility, severity or hospitalization risk, or on URI and influenza infection risk (IVW P≫0.05; Table 2). Although we are underpowered to detect the causal effects sizes that we observe, we are well-powered to detect more modest causal effects of sleep fragmentation on our outcomes. For instance, at 80% power, we are sufficiently powered to detect causal ORs of >1.37, >1.10 and >1.23 for COVID-19 severity, susceptibility and hospitalization risk, respectively, and >1.49 and >1.23 for influenza and URI. We can therefore effectively rule out causal effects larger than these of sleep fragmentation on our respiratory infection outcomes.

To demonstrate the robustness of the insomnia findings, we performed sensitivity analysis using 48 genetic variants robustly associated with insomnia in the UK Biobank cohort^4^. Despite the dramatically lower statistical power to detect a causal effect, we were still able to see the impact of insomnia on both COVID-19 hospitalization (IVW OR = 1.13 [1.03-1.24], P=0.011) and URI (IVW OR = 1.11 [1.02-1.20], P = 0.013) and saw no strong evidence of pleiotropy (MR Egger intercept P>0.05). The effect sizes were much-attenuated in comparison to the larger set of insomnia instruments (Supp Table 3). No significant association was seen for COVID severity or influenza susceptibility (IVW P>0.05).

Rows with results in bold font are statistically significant after Bonferroni correction and those in italics are significant (IVW P<0.05) at the level of a single test but not after Bonferroni correction. NVar = number of exposure genetic instruments used.

## Discussion

Here we provide the first evidence that insomnia causally impacts the risk of developing respiratory infections including influenza and severe COVID-19. Leveraging the complex longitudinal health data from two large independent population-based cohorts, the UK Biobank and FinnGen, we assessed whether a prior diagnosis of insomnia led to an increased risk of either influenza or URI. Results from up to 23 years of follow-up diagnoses in both cohorts suggested a strong increase in URI and influenza infection risk for insomnia sufferers, though the increase in risk differed greatly between the two studies (e.g. influenza’s HR was 1.54 in UKB vs. 5.32 in FinnGen). This is likely due to FinnGen diagnoses consisting of only hospital records (both inpatient and outpatient) and therefore representing more severe cases of insomnia, URI and influenza, whereas diagnosis data from both hospital inpatient and primary care (doctor visits) were considered in the UK Biobank data.

We also used a framework from genetic epidemiology called Mendelian Randomization (MR), a form of instrumental variable analysis, that can estimate the causal association of an exposure on an outcome using genetic variants robustly associated with the exposure. We demonstrated that insomnia is strongly causally associated with an increased risk of URI, influenza, COVID-19 hospitalization and severity, and to a lesser extent, with an increased risk of initial SARS-CoV-2 infection. These novel findings are in line with earlier literature and together demonstrate the impact that sleep has on immune function, which then likely has a downstream effect on the ability to fight infection.

The earlier studies have systematically demonstrated that both subjective and objective measures of poor sleep, such as insufficient sleep and difficulty getting to sleep, are associated with increases in C-reactive protein (CRP) and cytokines like interleukin 1β (IL-1β) and interleukin 6 (IL-6), amongst others^9,18,31,32^. Furthermore, a study in 674 adults suggested an association with increased risk of respiratory infections^17^, though there is some evidence that the relationship is also likely to be bidirectional, with sleep changes also occurring as a result of viral infection^33,34^. In addition to the effect of unmeasured confounders such as shared environment and lifestyle being difficult to rule out, this has previously made causal inference difficult. Our findings are in line with these observations and go one step further in being able to assess causal, rather than correlative, associations showing that chronic insomnia symptoms are indeed a risk factor for infections.

Interestingly, we saw a stronger association with COVID-19 severity than with COVID-19 infection, despite having more power to detect association with COVID-19 infection than severity. We conjecture that this may be due to three factors. Firstly, insomnia may act on the severity of the respiratory infections more strongly than on the risk of initial infection, as seen with other risk factors like body mass index (BMI)^35–37^, obstructive sleep apnea (OSA)^38–40^, fasting blood glucose^41^, and high blood pressure^42,43^. This would be in line with evidence that those with severe COVID-19 have elevated levels of IL-6 and CRP when compared to non-severe COVID-19 patients^44^, given the relationship between chronic sleep disruption and higher levels of circulating inflammatory markers, but remains to be seen for other respiratory infections. Secondly, both insomnia and COVID-19 infection and severity are correlated with demographic measures and therefore not distributed uniformly in the population. For example, socioeconomic factors, occupation, age, sex and ethnicity are all associated with increased rates of insomnia and COVID-19 susceptibility and severity^2,45^. These uncaptured confounders are likely to affect the estimates from longitudinal analyses and, for inherited unmeasured factors, may result in pleiotropy in the causal estimates, more so as GWAS sample sizes increase^46^. For the insomnia exposure that used instruments from the most recent GWAS meta-analysis, there was some evidence of pleiotropy in the causal estimate on COVID-19 severity and hospitalization outcomes (Supp Table 3), though the sensitivity analyses using a more restricted set of instruments found no evidence of pleiotropy, albeit with more moderate causal effects on COVID-19 severity and hospitalization. Thirdly, we recognize that insomnia itself is a multifactorial disorder with a variety of potential causes and presentations, each of which may confer differing levels of risk for susceptibility to or severity of respiratory infections. Evidence suggests, for example, that short sleep (a common consequence of insomnia) and oxygen desaturation (a consequence of OSA, which could commonly be misdiagnosed as insomnia^47^) may both be independently associated with inflammation to varying degrees^6^. It is possible that different symptoms of insomnia have different downstream biological effects which, when considered separately, would show heterogeneous effects on susceptibility, but more homogenous effects on respiratory infection severity. We acknowledge that there is more work to be done to understand the specific causal mechanisms that drive the increase in infection risk and severity in those suffering from insomnia.

We note the following limitations, welcome feedback from the scientific community and will aim to address some of these issues in future studies in this line of research. First of all, FinnGen endpoints were identified using hospital records only, and consequently results of survival analyses are biased towards more serious cases of the prior (insomnia) endpoint. In comparison, in the UK Biobank diagnoses were captured in hospital and primary care data. It is therefore possible that the differences in hazard ratio we see between the two cohorts represent a) greater statistical power due to more severe insomnia diagnosis, b) a real difference in effect of insomnia on more severe (FinnGen) and less severe (UK Biobank) respiratory infections, c) differences between the cohort demographics or d) a combination of these factors. Confounding this issue is that the UK Biobank suffers from ascertainment bias, resulting in a cohort that is, on average, healthier and less socioeconomically deprived than the general UK population^48^ and it is likely, though not yet reported, that the FinnGen study also suffers from this bias. The insomnia prevalence that we observed (0.9% in FinnGen and 3.8% in UKB; Supp Table 5) point to low ascertainment in these cohorts compared to estimates of insomnia prevalence (7.9% in the UK^49^ and up to 11.7% in Finland^50^). While ascertainment bias has been shown to exaggerate the effects of a prior endpoint on survival time^51^, it is not currently known how this ascertainment bias might affect endpoint-to-endpoint survival analyses where both endpoints may be subject to this bias.

Secondly, while Mendelian randomization is a powerful tool to estimate causality, there were some noticeable limitations to its use in this study. Primarily, we could not produce easily interpretable causal estimate effect sizes due to both exposure and outcomes being binary phenotypes. Additionally, we selected our insomnia instruments from the largest GWAS meta-analysis, to date, of self-report insomnia in order to maximize statistical power and there was some evidence of horizontal pleiotropy in these variants as evidenced by the non-zero MR-Egger intercept (Supp Table 3). It is possible that, due to the large sample size of the meta-analysis and thus the high power to detect genetic associations, that a proportion of the selected instruments may be secondary associations for insomnia, being associated with a phenotype that itself influences insomnia risk. For instance, genetic correlation analyses identified a large overlap with depression which could be a large contributor to the overall pleiotropy of these instruments. We demonstrated that a strong causal association persisted even when restricting to a smaller set of non-pleiotropic variants, such as those identified in a UK Biobank-only GWAS of insomnia symptoms, though with a more modest causal effect. Furthermore, we attempted to assess specific sleep behaviors comorbid with insomnia, to understand whether specific features of poor sleep were driving the causal associations we saw. Unfortunately, due to the limited explanatory power of the available instruments, we were generally underpowered to detect all but the largest effect sizes in the current MR framework and so cannot yet attribute the risk to any specific feature of insomnia or poor sleep.

Thirdly, we were unable to provide longitudinal estimates for COVID-19 infection and severity in the context of a prior insomnia diagnosis, which would have better contextualized the MR findings. In both cohorts, the available health records did not overlap the pandemic period (beginning March 2020) and so there was no contemporaneous non COVID-19 diagnosis data, meaning that survival analyses were not appropriate. In simple logistic regression analyses, there was no evidence of enrichment of COVID-19 infections amongst those previously diagnosed with insomnia (OR=1.21 in UKB and OR=1.24 in FinnGen, both P>0.05), but the relative rarity of COVID-19 diagnoses in both cohorts (0.3% for UKB and 1.1% for FinnGen; Supp Table 5), however, limits the power to observe any associations and, therefore, the utility of these estimates.

## Methods

### Cohorts

FinnGen is a study of a population-based cohort of Finnish residents, from newborn to age 104 at baseline recruitment, that have consented to participate in regional biobanks in Finland. The study combines genetic data with electronic health record data derived from primary care registers, hospital in- and out-patient visits and prescription information and aims to understand the genetic etiology of, and drive the development of drugs to treat, a wide variety of diseases and disorders. The current release (R7) contains health and genetic data on up to 327,625 participants, primarily of Finnish ancestry. Diagnosis data extraction from the public healthcare records is ongoing with this data being released on a regular basis and all participants continue to be followed up unless they die (follow-up ends at death) or they withdraw their consent from the study (their data is removed entirely and is not used in analyses). When a study participant is recruited, their entire medical record is linked into the FinnGen database and not just subsequent healthcare provider visits, allowing a detailed understanding of their medical history.

The UK Biobank is a prospective study of over 500,000 participants, aged between 37 and 73 at recruitment, from the mainland UK population^52^. Participants were invited to undertake a baseline interview between 2006 and 2010 where data was collected on a variety of health and lifestyle measures, and where blood and urine samples were also taken for genetic and biochemistry analysis. Electronic health records, consisting of Hospital Episode Statistics in-patient (HES; max. N=440,512) and primary care (GP; max. N=231,364) were later linked up to provide longitudinal data on disease diagnosis, operations, medications, and deaths. Recent diagnoses are frequently added to the records and, as with FinnGen, the participants’ entire medical history is made available, and they continue to be followed-up unless they die or withdraw their consent. While the UK Biobank participants are drawn from the general UK population, the study suffers from recruitment bias and as such participants are on average healthier, more educated and less deprived than the average UK resident^48^.

### Phenotype/endpoint definitions

Diagnosis data in the FinnGen study is compiled from multiple sources, some of which are disease-specific, such as the cancer, visual impairment, kidney disease, and infectious disease registries, whilst others cover more general health visits such as the Primary Care (“Avohilmo”), Inpatient (“Hilmo”) and the Finnish Social Insurance Institution (KELA drug-purchase registers. Inpatient diagnoses and death records were available from 1969, whereas hospital outpatient procedures were available from 1998. For insomnia, upper respiratory infection (URI), and influenza endpoints we used the pre-existing FinnGen endpoint definitions, which utilize inpatient, outpatient, and death records to determine endpoint cases and controls. Study participants were classed as endpoint cases if they had at least one record with a relevant ICD-8, ICD-9, or three-digit ICD-10 code assigned to it. The relevant codes were:

- insomnia: “F51.0”, “G47.0” (ICD-10)
- upper respiratory infection (URI): “J06”, “J06.0”, “J06.9” (ICD-10) or “465” (ICD-9 and ICD-8)
- influenza: “J09”, “J10”, “J10.0”, “J10.1”, “J10.2”, “J10.8”, “J11”, “J11.0”, “J11.1”, “J11.2”,“J11.8” (ICD-10) or “487” (ICD-9) or “470”, “471”, “472”, “473”, “474” (ICD-8).

If the ICD codes listed above contained subcodes, then these were included in the endpoint diagnosis definition. For instance, including ICD-9 code “465” in the URI endpoint meant that we included the subcodes “465.0”, “465.8” and “465.9” in addition to the original “465” code. For the purposes of the survival analysis in which diagnosis date is used, the date of diagnosis was the first date at which the participant was diagnosed with any of the included codes. Control groups were created separately for each endpoint and were defined as those with no relevant diagnosis for the specific endpoint. Of 327,528 FinnGen participants, there were 2,834, 28,462 and 6,933 with insomnia, URI and influenza endpoints respectively (Supp Table 5).

The UK Biobank cohort has both self-report and EHR-based disease definitions. To define equivalent endpoints in the UK Biobank, we used only diagnostic information from the EHR data. Electronic health data from multiple sources has been linked to the UK Biobank to date. Currently, these include death, cancer, hospital inpatient, primary care (GP) diagnosis, primary care prescription and primary care registration records. To define insomnia and respiratory infection endpoints in the UK Biobank, we used only the hospital inpatient (HES; field 41234) and primary care diagnosis records (field 42040). In the hospital inpatient data, we included individuals as a case for the endpoint if they had at least one of the same ICD-10 or ICD-9 diagnosis codes used for FinnGen (see above) and, as with FinnGen, included participants with subcodes in the endpoint. In the primary care data, diagnoses were coded using the NHS-specific Read v2 or CTV3 codes. We used the following Read codes to define the respective endpoints:

- Insomnia: “1B1B0”, “1B1B1”, “1B1B2”, “E2742”, “Eu510”, “Fy00.”, “R0052”, “X007s”, “X007u”, “X76AF”, “X76AG”, “Xa7wV”, “XaIv5”, “XE1Yg”, “XE2Pv” (Read CVT3) or “Eu510”, “Fy00.”, “R0051”, “R0052” (Read v2)
- URI: “H0…”, “H050.”, “H05z.”, “H0z..”, “X1003”, “Xa1sb”, “XaDcC”, “XE0Xq” (Read CVT3) or “H0…”, “H050.”, “H05z.”, “H0z..”, “X1003” (Read v2)
- influenza: “H2…”, “H27..”, “H270.”, “H2700”, “H270z”, “H271.”, “H2710”, “H2711”, “H271z”, “H27y.”, “H27y1”, “H27z.”, “H2y..”, “H2z..”, “XaQQp”, “XE0YK”, “XM0rz” (Read CVT3) or “H2…”, “H27..”, “H270.”, “H2700”, “H270z”, “H271.”, “H2710”, “H2711”, “H271z”, “H27y.”, “H27y1”, “H27z.” (Read v2)

As with FinnGen, date of diagnosis for an endpoint was taken as the date of the first identified visit with any of the included ICD-9, ICD-10, Read v2 or Read CTV3 codes and so the first diagnosis could be either a hospital inpatient or primary care visit. As primary care data is only available in a subset of participants, unlike hospital inpatient data, we limited endpoint definition and thus subsequent analyses to those with both hospital inpatient and primary care data. Of 231,364 participants with both HES and GP records available, there were 8,693, 55,250 and 12,948 with diagnoses of insomnia, URI and influenza, respectively, in the UK Biobank (Supp Table 5).

### COVID-19 diagnoses

Diagnoses of SARS-CoV-2 infection (COVID-19) in Finland are recorded in the THL (Finnish Institute for Health and Welfare) Infectious Disease Register, from which the COVID-19 diagnoses have been extracted and linked to FinnGen participants. In release 7 of FinnGen, diagnoses were available until 2021/05/27, at which point there were 3,497 unique individuals with a positive lab-confirmed COVID-19 diagnosis. Diagnoses were primarily by PCR (N=3,424), with a small proportion of samples diagnosed through antigen testing (N=9), or antibody testing (N=6), and 57 samples with a missing diagnosis type.

In the UK Biobank, COVID-19 diagnosis data was obtained from data field 40100. This field is derived using linked data collected by Public Health England (PHE), Public Health Scotland (PHS) and SAIL for England, Scotland and Wales, respectively. We used diagnosis data with a cut off of 2020/10/02 and had data on 1,713 unique samples with a positive COVID-19 diagnosis, of which 733 had both HES and GP data available. All samples included in UKB data field 40100 were diagnosed through PCR testing (https://biobank.ndph.ox.ac.uk/ukb/exinfo.cgi?src=COVID19).

### Genetic Data and Analyses

To perform the Mendelian randomization analyses for the influenza and URI outcomes, we used summary statistics generated by the FinnGen release 7 (R7) GWAS pipeline for the “J10_INFLUENZA” and “J10_ACUTEUPPERINFEC” endpoints respectively. Samples were genotyped using Illumina and Affymetrix chip arrays (Illumina Inc., San Diego, and Thermo Fisher Scientific, Santa Clara, CA, USA) and imputed to GRCh38/hg38 using Beagle v4.1^53^ with the SISu v3.0 reference panel, consisting of 3,775 whole genome-sequenced Finnish individuals^54^ (see https://dx.doi.org/10.17504/protocols.io.nmndc5e for the complete imputation and QC protocol). A total of 16,692,023 imputed genotypes were available in 309,154 participants. In the GWAS of influenza, there were 6,692 cases and 261,719 controls whereas for the URI GWAS there were 27,468 cases and 255,610 controls. These GWA analyses were performed using REGENIE^55^ v2.0.2 and in the model-building step (step 1) were adjusted for age at follow-up end (2019/12/31) or death, sex, genotyping batch and the first 10 genetic principal components. Step 1 was performed using leave-one-chromosome-out (LOCO) prediction with 55,139 well-imputed (imputation INFO > 0.95 in all batches) common (MAF > 1%) genetic variants with <3% missingness that had been LD pruned using a 1Mb window and r^2^ threshold of 0.1.

### Survival Analyses

We performed endpoint-to-endpoint survival analyses, which compare the risk of developing an outcome endpoint if subject to diagnosis of a prior endpoint and accounting for the time taken to be diagnosed with the outcome. We followed a near-identical approach to the FinnGen Risteys pipeline (see “Survival analyses between endpoints” at https://risteys.finngen.fi/documentation), only considering the date of first diagnosis for each endpoint. Unlike in the Risteys FinnGen pipeline we did not follow a case-cohort design, which involves selecting all cases and a fixed number of controls through random sampling; in both the FinnGen and UK Biobank survival analyses, we used all available controls. We performed this analysis using the Python module “lifelines” (v0.26.0)^56^ with Python (v3.8.11 for FinnGen, v3.7.11 for UK Biobank).

Briefly, study start and end dates were chosen in each study based on the availability of records for the majority of participants (see below). Participants who were prevalent cases of the outcome endpoint (those with an outcome diagnosis before the study start date) were removed (see Supp Table 6 for sample exclusion counts). Prior endpoint cases whose first diagnosis occurred before the study start were given a diagnosis date of the study start date. Prior endpoint cases whose first diagnosis occurred after the study start date were separated into two entries corresponding to their time as controls (from date of study entry to diagnosis date) and as cases (from diagnosis date to date of study exit). These individuals are each treated as two separate participants, the control who “leaves” the study on the diagnosis date and the case who “enters” the study on the diagnosis date.

The survival model used in this analysis can be written as:

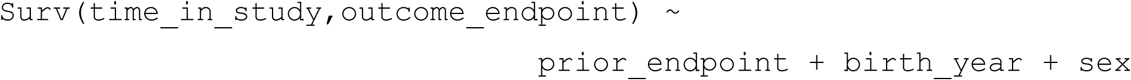

where “prior_endpoint” and “outcome_endpoint” were binary variables representing their case-control status and “time_in_study” was calculated from date of study entry to date of study exit. For sensitivity analyses, BMI was added as an extra term in the additive model and those without a BMI measurement were excluded in these analyses (exclusion counts provided in Supp Table 6). Date of study entry was taken as the study start date unless the participant was born later than this date (i.e., study entry is date of birth - this occurred only in FinnGen) or was diagnosed with the prior endpoint (their post-diagnosis or “case” record has study entry date as the date of diagnosis). Participants in both studies remain in the study and continue to accumulate diagnosis data unless they either withdraw their consent (and so are removed from the study entirely) or die. Therefore, for all included participants in FinnGen and the UK Biobank, the date of leaving the study was the specified study end date, unless the participant died before this date (study exit is the date of death) or was diagnosed with the prior endpoint (their pre-diagnosis or “control” record has study exit date as the date of diagnosis). For FinnGen (release 7), study start and end dates were set as 1998/01/01 and 2019/12/31, respectively, as the dates from which inpatient, outpatient and death records were available from and to for all participants. In the UK Biobank, the GP data is maintained in four distinct databases by three providers (see https://biobank.ndph.ox.ac.uk/showcase/showcase/docs/primary_care_data.pdf). To minimize the bias in UK Biobank-based analyses, we calculated a median primary care registration date (field 42038) in each database and selected a follow-up start date of 2002/03/01, the latest of these four median dates, ensuring that the majority of participants were already registered in each of the four databases. The study end date was identified as 2019/08/18 for the UK Biobank, the date of the latest available record from the primary care data (at the time of analysis). Diagnosis events in the UK Biobank hospital records that occurred later than this date were ignored. In the UK Biobank, some individuals had valid endpoint diagnoses but no valid date of diagnosis. For each prior and outcome endpoint pair, if either or both the endpoint diagnosis dates were invalid, the participant was removed from the analysis.

### Logistic Regression

These analyses were used to test whether insomnia diagnoses were enriched in participants with each of the outcome endpoints (URI, influenza, and COVID-19), regardless of which occurred first. The model we applied can be formulated as:

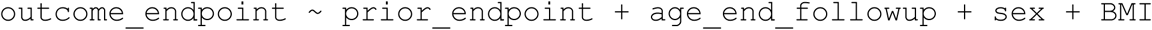

where “prior_endpoint” and “outcome_endpoint” were binary variables representing their case-control status for these endpoints. Unlike the survival analyses, we did not impose a follow-up start time and thus made no sample exclusions based on the date of outcome endpoint diagnosis (including those with no valid diagnosis date). It was still necessary to impose a follow-up end date, as the different registries contained within both FinnGen and UK Biobank were right-censored at different dates. Therefore, as with the survival analysis, for all endpoints except COVID-19 infection we imposed cut-off dates of 2019/12/31 and 2019/08/18 for FinnGen and UKB, respectively. Study participants with their first non-COVID endpoint diagnoses occurring after these dates were treated as controls for those endpoints. The COVID-19 diagnosis information was obtained from additional linked datasets for both cohorts and because all COVID-19 diagnoses occurred after the follow-up cut-off dates, we did not right-censor this outcome endpoint. In all logistic regression analyses, except with the COVID-19 outcome, the “age_end_followup” was defined as the participant’s age at the study’s respective cut-off date or their age at death if they died before this date. With the COVID-19 outcome, all participants who died before reaching the study’s cut-off date were excluded and so the “age_end_followup” simply represented each participant’s age at the cut-off date for all included samples. In performing this logistic regression analysis with the COVID-19 outcome, we did not consider death records after each study’s follow-up cut-off date and therefore made the assumption that study participants who survived until the study cut-off date also survived to the end of the COVID-19 diagnosis records (2021/05/27 in FinnGen and 2020/10/02 in the UK Biobank) or that cases and controls for the insomnia prior died at an equal rate after the follow-up cut-off date.

### Mendelian randomization

Mendelian randomization (MR) is an analysis method by which genetic variants robustly associated with an exposure (through GWAS) can be used to infer the one-directional causal impact of the exposure on an outcome by looking at the effect of the exposure’s variants on the outcome^57^. The causal effect is estimated from the gradient of the best-fit regression line of the variants’ effects on the outcome against their effect on the exposure. Evidence of a causal effect is assumed if the regression slope is significantly different from 0 (representing complete independence between the effects on the exposure and the outcome). Multiple methods exist for performing this regression, each with their own strengths and weaknesses^58^ and with different methods allowing for various degrees of violation of the core MR assumptions^59^.

Two-sample Mendelian randomization was performed in R (v3.6.3) using the package *TwoSampleMR*^*25,26*^ (v0.5.6). For our exposures, we used summary statistics from the most recent GWAS of insomnia in over 2.3 million individuals^27^ and from the largest GWAS of habitual short sleep in ∼410,000 UK Biobank participants^29^. To avoid sample overlap in our two-sample design, we used GWAS summary statistics from FinnGen (release 7) for the influenza and URI outcomes. With the COVID-19 outcomes, we obtained summary statistics of the GWAS meta-analyses^28^ that excluded both FinnGen and 23andMe for the B2 (“hospitalized” COVID-19 vs. population controls) and C2 (COVID-19 infection vs. population controls) phenotypes to avoid sample overlap.

As exposure instruments, we selected all variants with association P≤5×10^−8^ in the discovery GWAS and used the same study for both instrument selection and effect size determination (see Supp Table 7). Ideally, we would have performed the MR in a three-sample setting^60^ by determining effect sizes from an GWAS in an independent cohort to that of the exposure discovery GWAS, as two-sample MR settings suffer from winner’s curse bias, which can bias the causal estimate towards the null^61^. Independent (non-overlapping) cohorts of comparable size with both genetic data and either insomnia or habitual sleep duration are simply not available, making a three-sample setting unfeasible for this study.

We performed MR power calculations using the web-based tool *mRnd*^*62*^, available at https://shiny.cnsgenomics.com/mRnd/, and selected the option “Binary outcome”, where we input the known variables: sample size, case proportion and exposure variance explained by the instruments. We set the alpha to 0.05 and input the causal MR odds ratios that we wished to calculate the power level for. The estimated power for a range of odds ratios, along with the parameters used for each exposure-outcome pair, are provided in Supp Table 4. To calculate the variance explained in the exposure by each instrument, we used a piece of code from another MR study of Alzheimer’s Disease^63^ where they estimated the variance explained to calculate instrument strength (via the F-statistic) and which is available as the second supplementary file of that paper. We provide the estimated variance explained in the exposure by each variant in the MR variant lists (Supp Table 7) and for each exposure, we calculated the total exposure variance explained by summing the variance explained by each exposure variant included for that outcome (also Supp Table 7). For the insomnia exposure from the latest GWAS meta-analysis^27^, we obtained the estimate of the total variance explained as approximately 1%, from private communication with the authors.

## Supporting information

Supplementary Table Descriptions

## Data Availability

Instructions for accessing the FinnGen R6 influenza ("J10_INFLUENZA") and URI ("J10_ACUTEUPPERINFEC") GWA summary data are provided at https://finngen.gitbook.io/documentation/data-download, with details about R7 made available in Q2 2022. Prior to this, the authors can be contacted to provide the R7 summary data. The scripts for performing the Mendelian randomization and endpoint-to-endpoint survival analysis will be made available with the published version of the manuscript and can be provided on reasonable request to the authors.

## Data Availability

Individual-level data can be accessed on successful application to FinnGen and the UK Biobank cohorts. For FinnGen, applications for individual-level data can be made through the Finnish Biobanks’ “FinnBB” portal (https://finbb.fi/) and summary GWA data, including for the influenza and URI phenotypes, can be accessed through the FinnGen website (https://www.finngen.fi/en/access_results). For the UK Biobank, applications for individual-level data can be made through the UK Biobank portal at https://www.ukbiobank.ac.uk/enable-your-research/apply-for-access.

The FinnGen R7 GWA summary statistics for influenza and URI will become available to researchers in Q2 2022 at https://r7.finngen.fi/. The scripts for performing the Mendelian randomization and endpoint-to-endpoint survival analysis will be made available with the published version of the manuscript.

## Ethics

### FinnGen

All FinnGen participants provided informed consent for biobank research based on the Finnish Biobank Act (FBA). Prior to the FBA coming into effect (September 2013), participants recruited into the individual research cohorts provided study-specific consent for research. These consent permissions were transferred to the Finnish biobanks, at the conception of FinnGen in August 2017, after approval by Fimea (the Finnish Medicines Agency), the National Supervisory Authority for Welfare and Health. The recruitment protocols followed the biobank protocols approved by Fimea. The Coordinating Ethics Committee of the Hospital District of Helsinki and Uusimaa (HUS) statement number for the FinnGen study is Nr HUS/990/2017.

The FinnGen study is approved by the Finnish Institute for Health and Welfare (THL) under permit numbers THL/2031/6.02.00/2017, THL/1101/5.05.00/2017, THL/341/6.02.00/2018, THL/2222/6.02.00/2018, THL/283/6.02.00/2019, THL/1721/5.05.00/2019,

THL/1524/5.05.00/2020, and THL/2364/14.02/2020, by the Digital and Population Data Service Agency (DVV) under permits VRK43431/2017-3, VRK/6909/2018-3, VRK/4415/2019-3, the Social Insurance Institution (KELA) under permits KELA 58/522/2017, KELA 131/522/2018, KELA 70/522/2019, KELA 98/522/2019, KELA 138/522/2019, KELA 2/522/2020, KELA 16/522/2020, the Finnish Social and Health Data Permit Authority (Findata) under permit THL/2364/14.02/2020 and by Statistics Finland (Tilastokeskus) under permits TK-53-1041-17 and TK/143/07.03.00/2020 (formerly TK-53-90-20).

For freeze (release) 7 of the FinnGen study, the biobank access decisions include: THL Biobank BB2017_55, BB2017_111, BB2018_19, BB_2018_34, BB_2018_67, BB2018_71, BB2019_7,BB2019_8, BB2019_26, BB2020_1, Finnish Red Cross Blood Service Biobank 7.12.2017, Helsinki Biobank HUS/359/2017, Auria Biobank AB17-5154 and amendment #1 (August 17 2020), Biobank Borealis of Northern Finland_2017_1013, Biobank of Eastern Finland 1186/2018 and amendment 22 § /2020, Finnish Clinical Biobank Tampere MH0004 and amendments (21.02.2020 & 06.10.2020), Central Finland Biobank 1-2017, and Terveystalo Biobank STB 2018001.

### UK Biobank

The UK Biobank has received approval as a Research Tissue Bank from the North West Multi-centre Research Ethics Committee (MREC) under MREC permits 11/NW/0382 (2011-2016), 16/NW/0274 (2016-2021) and 21/NW/0157 (2021-2026). Researchers with approved applications are covered by these permits and are not required to seek additional approval, except in specific cases (see section B7 of the UK Biobank Access Procedures document: https://www.ukbiobank.ac.uk/media/omtl1ie4/access-procedures-2011-1.pdf). All participants of the UK Biobank study provided consent, at the baseline visit, for their personal data and biological samples to be collected and stored for research purposes. Participants are given the option to withdraw their consent at any time; any samples that have withdrawn their consent at the time of analysis were excluded from this study. A print version of the electronic consent form is stored as UK Biobank Resource 100252.

## Acknowledgements

Funding for the FinnGen project is provided by two grants from Business Finland (HUS 4685/31/2016 and UH 4386/31/2016) and by the following industry partners: AbbVie Inc., AstraZeneca UK Ltd, Biogen MA Inc., Bristol Myers Squibb (and Celgene Corporation & Celgene International II Sàrl), Genentech Inc., Merck Sharp & Dohme Corp, Pfizer Inc., GlaxoSmithKline Intellectual Property Development Ltd., Sanofi US Services Inc., Maze Therapeutics Inc., Janssen Biotech Inc, Novartis AG, and Boehringer Ingelheim. We acknowledge the support and work of the individual biobanks that the FinnGen cohort comprises: Auria Biobank

(www.auria.fi/biopankki), THL Biobank (www.thl.fi/biobank), Helsinki Biobank (www.helsinginbiopankki.fi), Biobank Borealis of Northern Finland (https://www.ppshp.fi/Tutkimus-ja-opetus/Biopankki/Pages/Biobank-Borealis-briefly-in-English.aspx), Finnish Clinical Biobank Tampere (www.tays.fi/en-US/Research_and_development/Finnish_Clinical_Biobank_Tampere), Biobank of Eastern Finland (www.ita-suomenbiopankki.fi/en), Central Finland Biobank (www.ksshp.fi/fi-FI/Potilaalle/Biopankki), Finnish Red Cross Blood Service Biobank (www.veripalvelu.fi/verenluovutus/biopankkitoiminta) and Terveystalo Biobank (www.terveystalo.com/fi/Yritystietoa/Terveystalo-Biopankki/Biopankki/). All Finnish biobanks are members of the European Biobanking and BioMolecular Resources Research Infrastructure (BBMRI-ERIC; https://www.bbmri-eric.eu/), and the Finnish Biobank Cooperative (FINBB; https://finbb.fi/) is the coordinator of the BBMRI-ERIC operations in Finland covering all Finnish biobanks.

This research has been conducted using the UK Biobank Resource under Application Number 22627. The UK Biobank study is funded by the following organizations: Medical Research Council, Wellcome Trust, Department of Health, Scottish Government, the Welsh Assembly Government, British Heart Foundation, Cancer Research UK and the Northwest Regional Development Agency.

The COVID-19 Human Genetics Initiative (HGI) (https://www.covid19hg.org/) is an ongoing international effort to identify the genetic factors that predispose people to a greater risk of COVID-19 (SARS-CoV-2 infection), severe COVID-19 and hospitalization from COVID-19. We would like to thank the COVID-19 HGI for publicly sharing their results ahead of publication, allowing us to investigate the causal effects of poor sleep on COVID-19.

Finally, we wish to acknowledge the participants of the UK Biobank and FinnGen studies for contributing to such important resources and for making work such as this possible.

