## Supplementary Table Descriptions for "The public health impact of poor sleep on severe COVID-19, influenza and upper respiratory infections"

### Supplementary Tables

- Supp Table 1: Results of endpoint-to-endpoint survival analyses in the combined UK Biobank HES and GP records and in FinnGen release 7 with insomnia as the prior endpoint and influenza and URI as the outcome endpoints. The model used is described in the methods section and was performed with (“Incl. BMI”) and without (“Excl. BMI”) BMI as a covariate. CI = 95% confidence interval.
- Supp Table 2: Results of the logistic regression analyses in both the UK Biobank and FinnGen release 7 for insomnia versus influenza, URI and COVID-19. The logistic regression model used and follow-up cut off dates are described in the methods section.
- Supp Table 3: Mendelian randomization results for four exposures: insomnia (Watanabe), frequent insomnia (Lane), short sleep (Dashti) and number of sleep episodes (Jones) and five outcomes: severe COVID, hospitalized COVID, COVID infection (all COVID HGI), upper respiratory infection and influenza (both FinnGen). Three methods were used - inverse variance weighted (IVW) MR, weighted median MR and MR Egger. NVar = number of genetic variants used as instruments for the exposure vs. the specific outcome.
- Supp Table 4: Power calculations for MR analyses of the four exposures and five outcomes. For the specific odds ratios, we calculated the power that we had in each analysis to identify odds ratios of that size. For the specific power thresholds, we calculated the minimum odds ratio that we would have that specific power to detect in each analysis.
- Supp Table 5: Cohort-specific demographic information for samples included in the survival and logistic regression analyses. All numbers were calculated in the subset of individuals in each cohort with both sex and diagnosis data available. For the UK Biobank, only those with both hospital episode statistics (HES) data and primary care (GP) data were included in these analyses.
- Supp Table 6: Sample inclusion and exclusion counts for all logistic regression (“LogReg”) and survival (“Surv”) analyses, both including BMI (“incl. BMI”) and excluding (“excl. BMI”). In analyses where more than one exclusion criterion was used (i.e. “Surv incl. BMI” or the COVID-outcome “LogReg incl. BMI” analyses), samples were first excluded based on missing BMI, then secondly if they had a missing diagnosis date for the exposure followed by those with a missing diagnosis date for the outcome (UK Biobank Surv only), then thirdly if the outcome diagnosis occurred before the follow-up start date (Surv only) and participants who died before the end of follow-up (COVID-19 analyses only). A missing value in the sample exclusion counts columns indicates that the specific exclusion was not made for that analysis.

- Supp Table 7: MR variant list detailing which variants were used in which analysis and their exposure association summary statistics. “Insomnia (frequent)” refers to instruments identified in the 2019 Lane et. al. [https://doi.org/10.1038/s41588-019-0361-7] study of frequent insomnia symptoms in the UK Biobank whereas “Insomnia (meta-analysis)” refers to instruments identified in the latest GWAS meta-analysis of insomnia symptoms [https://doi.org/10.1101/2020.12.07.20245209]. The (Pseudo)-R2 column was calculated using the script provided as Supp File 2 in a causal study of periodontitis and Alzheimer’s Disease [https://doi.org/10.1371/journal.pone.0228206].
